# The developmental trajectory of EEG alpha coherence in autistic toddlers with and without language delay

**DOI:** 10.64898/2026.06.03.26354124

**Authors:** Sophie Mandl, Haerin Chung, Winko W. An, Rebecca P. Thomas, Anjali Bose, Susan Faja, Carol L. Wilkinson

## Abstract

Although language acquisition delays are frequently observed in children with autism spectrum disorder (autism), our current understanding of the neurobiological mechanisms underlying language development in autism is sparse. Previous studies have found resting-state electroencephalography (EEG) power to be associated with language abilities in autistic children. However, longitudinal studies examining resting-state EEG phase coherence in relation to language development in preschool-aged children with autism are limited. This study aimed to characterize age- and group-related changes in whole-brain coherence in neurotypical children and in autistic children with and without language delay. Resting-state EEG and language data were collected at 2, 3, and 4 years of age. Peak phase coherence within the alpha band (6-11 Hz) was calculated at each timepoint and differences in the developmental trajectory of peak alpha coherence (PAC) were analyzed. In neurotypical children, PAC increased between 2 and 4 years of age. In contrast, PAC did not significantly change with age in children with autism. However, when examining autistic children based on language delay status, PAC increased with age in autistic children without language delay, but not in children with language delay. Exploratory analysis revealed evidence for an interaction between PAC and age, suggesting that the direction of the association between PAC and VDQ varied across age. Overall, these results support previous findings of altered oscillatory connectivity in autism and suggest that differences become apparent early in development. Importantly, phase coherence may not only differentiate diagnostic groups but also capture meaningful variability *within* the autism group. Future research should further investigate the use of EEG coherence as a biomarker of language development in autism.

**Lay summary:** Children with autism often experience delays in language development, but the brain mechanisms behind this are not well understood. This study tracked brain activity in children with and without autism from ages 2 to 4 years and found that a measure of how well different brain regions communicate with each other, called phase coherence, increased over time in typically developing children and in autistic children without language delays, but not in autistic children with language delays. These findings suggest that this brain measure could help identify which autistic children are at risk for language difficulties early in development.

## Introduction

Autism spectrum disorder (autism) is a prevalent neurodevelopmental condition that is characterized by challenges in social communication and presence of restricted and repetitive behavior. Moreover, delays in language acquisition are common in children with autism. More than 50% of children diagnosed under three years of age exhibit language delays, and approximately 30% of children continue to be minimally verbal throughout their life (Eigsti et al., 2011; Tager-Flusberg & Kasari, 2013). However, there is substantial heterogeneity both in children’s initial language presentation and in their later language development, with approximately 25% of children diagnosed with autism ultimately developing language abilities consistent with chronological age expectations (Anderson et al., 2007; Delehanty et al., 2018). Even though language impairment is no longer required for a diagnosis of autism, language functioning by school age is one of the best predictors of later academic performance, emotional regulation, and behavioral functioning (Botting et al., 2016; Conti-Ramsden et al., 2013).

Given that language functioning carries such significant long-term implications, understanding its neural basis in autism becomes critical. Language is a highly complex ability that relies on the coordinated activity of widely distributed brain networks. Disruptions to the development and coordination of these networks are therefore a plausible mechanism underlying the language difficulties commonly observed in autism. Resting-state EEG is well-suited to capturing such disruptions: the recorded oscillatory activity reflects rhythmic fluctuations in neural excitability, which are widely interpreted as a marker of coordinated neural communication. Among the frequency bands observable in EEG, alpha oscillations are of particular interest, as they have been linked to cortical inhibition, attentional gating, and long-range synchrony, all of which are fundamental to language processing (Bastiaansen & Hagoort, 2006; Foxe & Snyder, 2011; Klimesch et al., 2007; Palva & Palva, 2011). Moreover, alpha activity is typically strong, consistent, and easy to detect during relaxed wakefulness, which makes it a robust metric for between-subject analyses.

EEG power spectral analysis is the most widely used approach to quantify resting-state EEG (Neo et al., 2023). In autism, resting-state EEG studies have overwhelmingly found reduced absolute and relative alpha power across development, from early childhood (Luschekina et al., 2017; Machado et al., 2015; Pierce et al., 2021) to adolescence and adulthood (Dawson et al., 1995; Murias et al., 2007). There is also evidence that resting-state EEG spectral power is linked to language development in autistic children. For example, Wilkinson et al. (2019) found resting frontal gamma power to correlate negatively with expressive language in high-likelihood two-year-olds (i.e. children with autistic siblings). Similar results were observed in an older sample of two-to five-year old autistic children, with reduced frontal gamma power being significantly associated with higher expressive language skills and non-verbal cognitive abilities (Mukerji et al., 2025). Extending this work, one recent study showed that both aperiodic and periodic features of the EEG power spectrum are differentially related to language abilities in autism. In particular, lower aperiodic offset and higher periodic alpha power were associated with better expressive and receptive language skills, and autistic children with language impairment exhibited reduced periodic alpha power compared to those without language difficulties (Chen et al., 2026).

However, spectral power, while informative, is a restricted measure of brain activity because it only captures the strength of neural oscillations, not the degree of integration between brain regions. By contrast, phase coherence measures the consistency of phase relationships between two neural signals over time, offering insight into the functional connectivity of distributed networks that support complex cognition and behavior. This distinction is particularly important in autism research, where it is hypothesized that altered connectivity may underlie some of the core cognitive and language deficits seen in autism (Courchesne & Pierce, 2005; Just et al., 2004). Existing research has predominantly reported reduced coherence in lower frequency bands, including alpha, and increased coherence at higher frequencies (O’Reilly et al., 2017; Ye et al., 2014). Notably, differences in alpha coherence between autism and neurotypical individuals appear to be emergent, with studies in high-likelihood infants reporting hyperconnectivity (Orekhova et al., 2014) and studies in autistic children and adults finding hypoconnectivity (Barnes et al., 2025; Boersma et al., 2013; Coben et al., 2008; Murias et al., 2007).

Despite growing evidence on the neuronal underpinnings of autism, several major gaps remain in the literature. First, there is a scarcity of longitudinal data on the developmental trajectory of functional connectivity in children diagnosed with autism. Most studies to date have cross-sectionally examined older children, adolescents, or adults. Yet the brain develops rapidly in the first years of life, with initial synaptogenesis supporting early connectivity between brain regions, followed by synaptic pruning refining neural networks to improve efficiency and functional specialization. It is therefore of interest to compare the early neurodevelopmental trajectory of children with autism to children without autism. Second, our current understanding of the neurobiological mechanisms underlying language deficits in autism is sparse, which has limited our ability to employ effective interventions and help children with autism reach their developmental potential. To the best of our knowledge, there is limited evidence on the development of alpha coherence across early childhood in autism, and particularly on whether language delay status shapes this neural trajectory over time.

Therefore, the aim of the present study was twofold. First, we examined whether there are differences in the developmental trajectory of peak alpha coherence (PAC) from 2 to 4 years of age between children with and without autism. Second, to capture differences between children within the autism group, we tested whether alterations in PAC, and its change over time, were associated with language delay status at 2, 3, and 4 years of age. We hypothesized that (1) autistic children would exhibit reduced PAC compared to age-matched controls from 2 to 4 years, and (2) autistic children with language delay (autism-LD) would show lower PAC than those without language delay (autism-noLD), with group differences becoming more pronounced with age.

## Methods

### 2.1. Participants

The present work combined data from one cross-sectional and three longitudinal studies conducted across four laboratories at Boston Children’s Hospital. All children were recruited from the greater Boston area between 2007 and 2024. Inclusion criteria were (a) English as their primary language, (b) no pre- or postnatal complications, and (c) no genetic or neurologic disorders. Together, the studies acquired data from 53 autistic and 72 neurotypical children at 2 years (*M_age_* = 27.09, *SD_age_* = 3.97 months), from 66 autistic and 95 neurotypical children at 3 years (*M_age_* = 39.40, *SD_age_* = 3.91 months), and from 56 autistic and 57 neurotypical children at 4 years of age (*M_age_* = 54.01, *SD_age_* = 3.43 months; see Table S1 in the Supplement for the number of participants from each study). All four studies were approved by the Boston Children’s Hospital institutional review board and conducted in accordance with the Helsinki Declaration of 1975. A parent or legal guardian gave informed written consent before each child’s inclusion.

#### 2.1.1. Brain Indicators of Developmental Growth (BRIDGE) study

This longitudinal study aimed to identify EEG features associated with language deficits in children aged 2 to 5.5 years old across multiple neurodevelopmental disorders, including autism, Fragile X Syndrome, and Trisomy 21 (Down syndrome). Participants were seen twice, one year apart. At both timepoints, participants completed a resting-state EEG measurement, parent-child interaction, as well as nonverbal and language assessments. All autistic participants received a clinical diagnosis from either a developmental behavioral pediatrician, neurologist, or child psychologist.

#### 2.1.2. Individual Development of Executive Attention (IDEA) study

This longitudinal study aimed to determine whether deficits in executive control in children with autism can be identified before preschool and assess whether executive control plays a role in development and maintenance of core autism features. Children were enrolled at either 2- or 4 years of age and followed with two additional follow-up visits that were scheduled one year apart. While language development was not a major focus of the original study, language abilities, parent-child interactions, and resting-state EEG were assessed at each timepoint. Autism diagnoses were established by a licensed psychologist using the Autism Diagnostic Observation Schedule, Second Edition (ADOS-2), and the Autism Diagnostic Interview, Revised (Lord et al., 2012; Lord et al., 1994).

#### 2.1.3. Infant Sibling Project (ISP) study

This longitudinal study followed infants with autistic siblings to identify infant EEG measures predictive of a later autism diagnosis or language impairment. Infants were enrolled between 3 and 12 months of age and those diagnosed with autism were seen at both 24 and 36 months, when they completed resting-state EEG, parent-child interaction, as well as a nonverbal and language assessment. Autism diagnosis was determined by a licensed clinician using DSM-5 criteria with review of ADOS-2, developmental assessments, and parent-child interactions.

#### 2.1.4. Sensory Processing Adaptation (SPA) study

This cross-sectional study aimed to identify EEG features associated with differences in sensory processing in 3- to 4-year-old children with autism. Participants were assessed with resting-state EEG, a nonverbal-, and a language examination. Participants received a community diagnosis of autism and met criteria for autism on the ADOS-2.

### 2.2. EEG data acquisition and preprocessing

Continuous, resting-state EEG data were collected for 2 to 5 minutes using either a 64-channel Geodesic Sensor Net System or a 128-channel HydroCel Geodesic Sensor Net (Electrical Geodesics, Inc., Eugene, OR) that was connected to a DC-coupled amplifier (Net Amps 200 or Net Amps 300, Electrical Geodesics, Inc.). Data were sampled at either 250 or 500 Hz and referenced to a single vertex electrode.

Raw EEG data were exported to MATLAB (version R2025a) and preprocessed using the Harvard Automated Preprocessing Pipeline for EEG, Version 3 (HAPPE v.3). All data were high-pass filtered at 1 Hz and low-pass filtered at 100 Hz. EEG data sampled at 500 Hz were resampled to 250 Hz. Artifacts were removed with 60 Hz line noise removal, bad channel rejection, and wavelet thresholding. In addition to electrodes in the 10-20 system, the following channels were selected in the 64-channel net: 14, 12, 7, 2, 1, 8, 3, 16, 9, 4, 58, 57, 20, 21, 5, 55, 53, 56, 25, 22, 18, 30, 43, 47, 50, 26, 33, 38, 41, 51, 32, 45, 36, 44; and in the 128-channel net: 26, 16, 2, 23, 3, 28, 19, 4, 117, 39, 41, 47, 57, 37, 42, 12, 6, 112, 7, 106, 55, 87, 93, 104, 115, 103, 98, 96, 100, 67, 77, 65, 69, 75, 89, 90. These 51 electrodes were chosen because they evenly cover the scalp (see Figure S1 in the Supplement). Data were then re-referenced to the average reference and divided into 1-second segments with 50% overlap. Segments were further evaluated using HAPPE’s amplitude and joint probability criteria to reject segments with retained artifacts. Participants were excluded from subsequent analyses if their EEG had fewer than 60 segments and/or less than 80% good channels. Based on these data quality metrics, six out of 125 participants were excluded in the 2-year cohort, one out of 161 was excluded in the 3-year cohort, and one out of 113 was excluded in the 4-year cohort.

### 2.3. Phase coherence calculation

To quantify functional connectivity between distributed brain regions, current source density (CSD) was first applied to the EEG data to improve spatial resolution and reduce volume conduction effects. Phase coherence was then computed for each possible pair of electrodes (n = 51 electrodes, yielding 1275 unique pairs in total) using spectral_connectivity_epochs in MNE-Python. Phase coherence captures the temporal consistency of phase differences between two oscillatory signals over time, reflecting the stability of their phase-locking relationship: values range from 0 (indicating random, uncorrelated phase distributions and thus no functional coupling) to 1 (indicating perfect phase alignment and maximal synchronization). To obtain a single index of whole-brain, or “global”, phase synchronization, coherence values were averaged across all 1275 pairs, yielding one mean coherence value per participant per frequency. The peak coherence value within the alpha frequency band (6-11 Hz) was then identified for each participant using the scipy find_peak function. This approach was chosen over band averaging to capture the most salient, participant-specific connectivity strength at the frequency of maximal alpha expression, which may shift developmentally or differ between autism and neurotypical groups.

### 2.4. Developmental language assessment

Language ability was determined at 2, 3, and 4 years of age with the Mullen Scales of Early Learning (MSEL), which is a standardized developmental assessment for children aged between 0 and 68 months (Mullen, 1995). Language delay status was based on the Expressive Language subscale, which evaluates verbal abilities through milestones such as babbling, single words, word combinations, and early grammar. Children were classified as language delayed (LD) at each age point if their expressive language standard scores fell more than 1.5 standard deviations below the normative mean (t-score < 37). For exploratory analyses, a verbal developmental quotient (VDQ) was computed as the mean of the age-equivalent scores from the Expressive and Receptive Language subscales, divided by chronological age and multiplied by 100. A nonverbal developmental quotient (NVDQ) was calculated in the same manner using the Visual Reception and Fine Motor subscales.

### 2.5. Statistical analyses

All statistical analyses and visualizations were conducted in Python (version 3.13.4) and R (version 4.5.1). Group differences in EEG quality metrics and MSEL scores were calculated using independent samples t-tests and differences in language delay status were computed with Fisher’s exact test. For Aim 1, the developmental trajectory of PAC was assessed using a linear mixed effects model (Python *statsmodels* package, *mixedlm*, version 0.14.6) with fixed effects for diagnostic group (autism, no autism), age, and their interaction (group x age). A random intercept for each participant was included to account for within-subject dependency arising from repeated observations. To assess longitudinal differences in PAC within the autism group based on language delay at 2, 3, and 4 years of age (Aim 2), a second linear mixed effects model with fixed effects for group (autism-LD, autism-noLD), age, and their interaction (group x age) was run. As each aim involved a single pre-specified hypothesis, the two confirmatory models were treated as independent tests and no correction for multiple comparisons was applied. All reported p-values are two-tailed, with the statistical significance level set to p < 0.05.

In addition to the confirmatory analyses, we fit an exploratory Bayesian mixed-effects model (R *brms* package, version 4.5.1) to examine whether there was an association between PAC and VDQ, and whether this link varied with age within the autism group. The model included participant-specific random intercepts, and predictors (PAC, age) were mean-centered prior to analysis.

## Results

### 3.1. Participant characteristics

The final analysis included 119 participants in the 2-year cohort, 160 participants in the 3-year cohort, and 112 participants in the 4-year cohort. The demographics of these participants are depicted in Table 1. Sex distribution was comparable between diagnostic groups at 2 years (χ^2^(1) = 2.38, p = .123) but differed significantly at 3 years (χ^2^(1) = 11.45, p = .001) and 4 years (χ^2^(1) = 6.32, p = .012), with the autistic group containing a higher proportion of males than the neurotypical group.

**Table 1.**
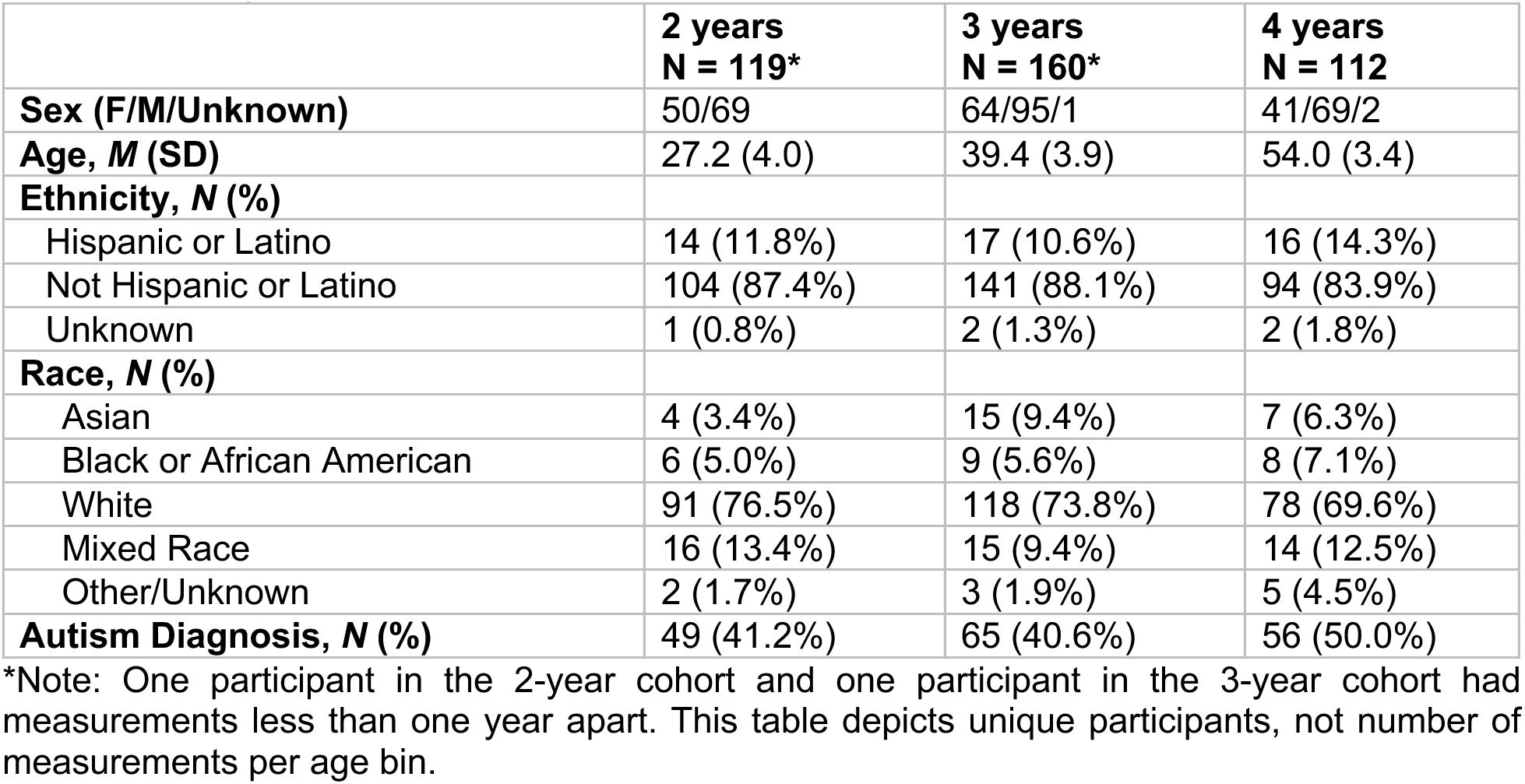
Participant characteristics.

|  | **2 years**<br>**N = 119\*** | **3 years**<br>**N = 160\*** | **4 years**<br>**N = 112** |
| --- | --- | --- | --- |
| **Sex (F/M/Unknown)** | 50/69 | 64/95/1 | 41/69/2 |
| **Age, *M* (SD)** | 27.2 (4.0) | 39.4 (3.9) | 54.0 (3.4) |
| **Ethnicity, *N* (%)** |  |  |  |
| Hispanic or Latino | 14 (11.8%) | 17 (10.6%) | 16 (14.3%) |
| Not Hispanic or Latino | 104 (87.4%) | 141 (88.1%) | 94 (83.9%) |
| Unknown | 1 (0.8%) | 2 (1.3%) | 2 (1.8%) |
| **Race, *N* (%)** |  |  |  |
| Asian | 4 (3.4%) | 15 (9.4%) | 7 (6.3%) |
| Black or African American | 6 (5.0%) | 9 (5.6%) | 8 (7.1%) |
| White | 91 (76.5%) | 118 (73.8%) | 78 (69.6%) |
| Mixed Race | 16 (13.4%) | 15 (9.4%) | 14 (12.5%) |
| Other/Unknown | 2 (1.7%) | 3 (1.9%) | 5 (4.5%) |
| **Autism Diagnosis, *N* (%)** | 49 (41.2%) | 65 (40.6%) | 56 (50.0%) |

\*Note: One participant in the 2-year cohort and one participant in the 3-year cohort had measurements less than one year apart. This table depicts unique participants, not number of measurements per age bin.

EEG data quality, MSEL language scores, and language delay status are summarized separately for participants with and without an autism diagnosis in Table 2.

**Table 2.**
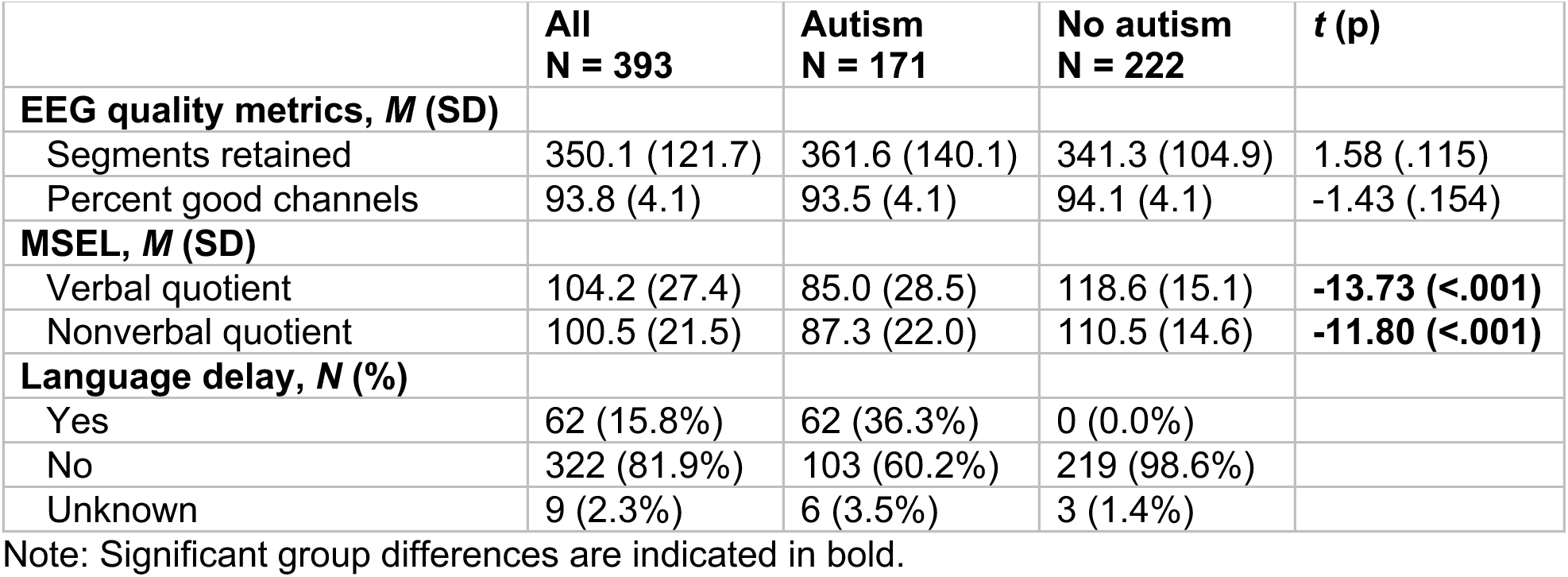
Group differences in EEG quality and language scores averaged across age bins.

|  | **All**<br>**N = 393** | **Autism**<br>**N = 171** | **No autism**<br>**N = 222** | ***t* (p)** |
| --- | --- | --- | --- | --- |
| **EEG quality metrics, *M* (SD)** |  |  |  |  |
| Segments retained | 350.1 (121.7) | 361.6 (140.1) | 341.3 (104.9) | 1.58 (.115) |
| Percent good channels | 93.8 (4.1) | 93.5 (4.1) | 94.1 (4.1) | -1.43 (.154) |
| **MSEL, *M* (SD)** |  |  |  |  |
| Verbal quotient | 104.2 (27.4) | 85.0 (28.5) | 118.6 (15.1) | **-13.73 (<.001)** |
| Nonverbal quotient | 100.5 (21.5) | 87.3 (22.0) | 110.5 (14.6) | **-11.80 (<.001)** |
| **Language delay, *N* (%)** |  |  |  |  |
| Yes | 62 (15.8%) | 62 (36.3%) | 0 (0.0%) |  |
| No | 322 (81.9%) | 103 (60.2%) | 219 (98.6%) |  |
| Unknown | 9 (2.3%) | 6 (3.5%) | 3 (1.4%) |  |

Note: Significant group differences are indicated in bold.

While there were no significant differences in EEG quality metrics, groups significantly differed in their VDQ (t = −13.74, p < .001) and NVDQ (t = −11.80, p < .001), as well as their language delay status (p <.001; Fig. 1).

**Fig. 1.**
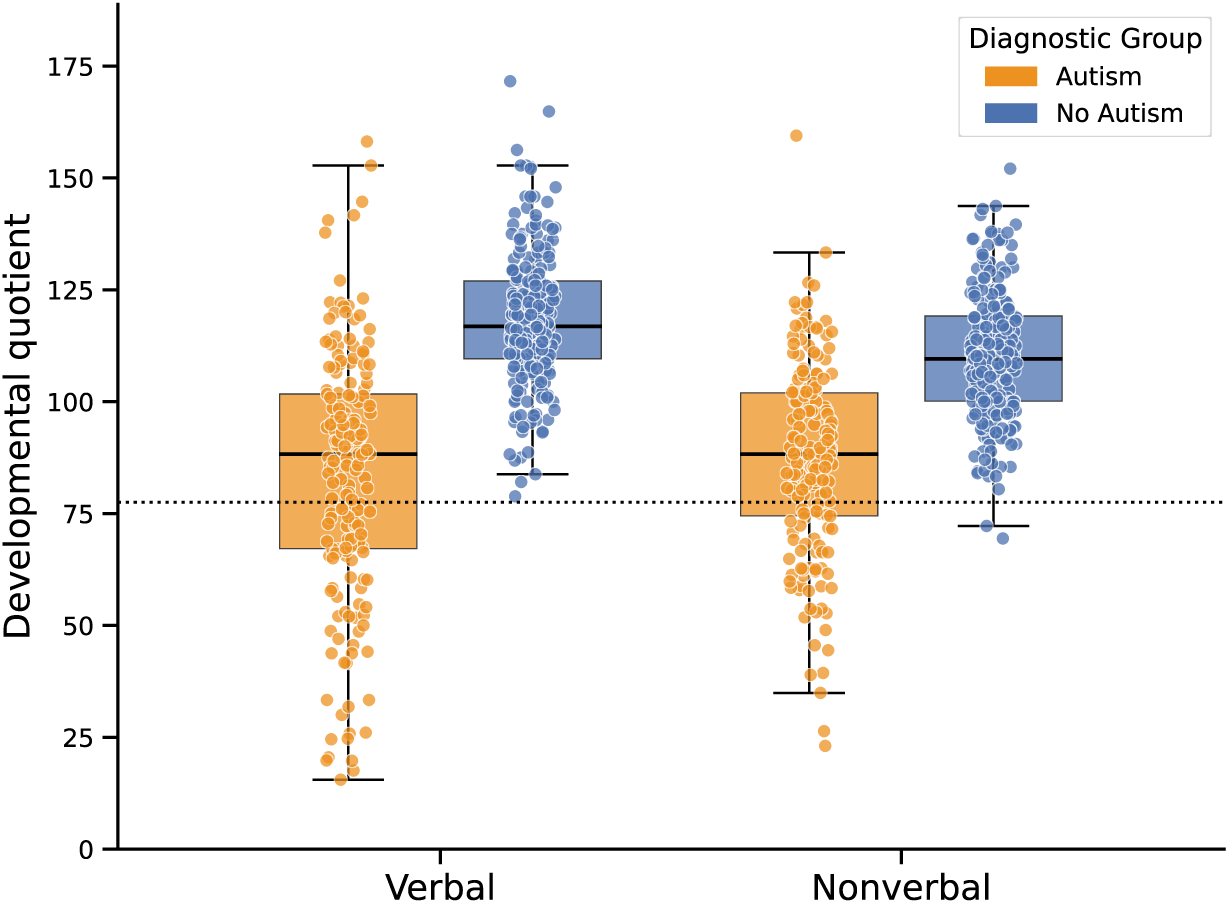
Differences in verbal and nonverbal developmental quotients between children with (orange) and without autism (blue). The dotted line indicates the language delay cut-off.

### 3.2. EEG alpha coherence in toddlers with and without autism

The first aim of the present study was to examine differences in the developmental trajectory of PAC from 2 to 4 years of age between children with and without autism. The final sample used for the coherence analyses comprised 171 children with autism and 221 typically developing children across all three age bins, as one participant did not have a valid alpha peak (see Figure S2 in the Supplement).

The linear mixed effects model revealed a significant fixed effect of age (β = 0.001, SE = 0.000, z = 4.82, p < .001) and a significant group x age interaction (β = −0.001, SE = 0.000, z = −2.37, p = .018). PAC increased significantly with age in children without autism (β = 0.001, SE = 0.000, z = 4.82, p < .001), but not in children with autism (β = 0.000, SE = 0.000, z = 1.35, p = .178; Fig. 2).

**Fig. 2.**
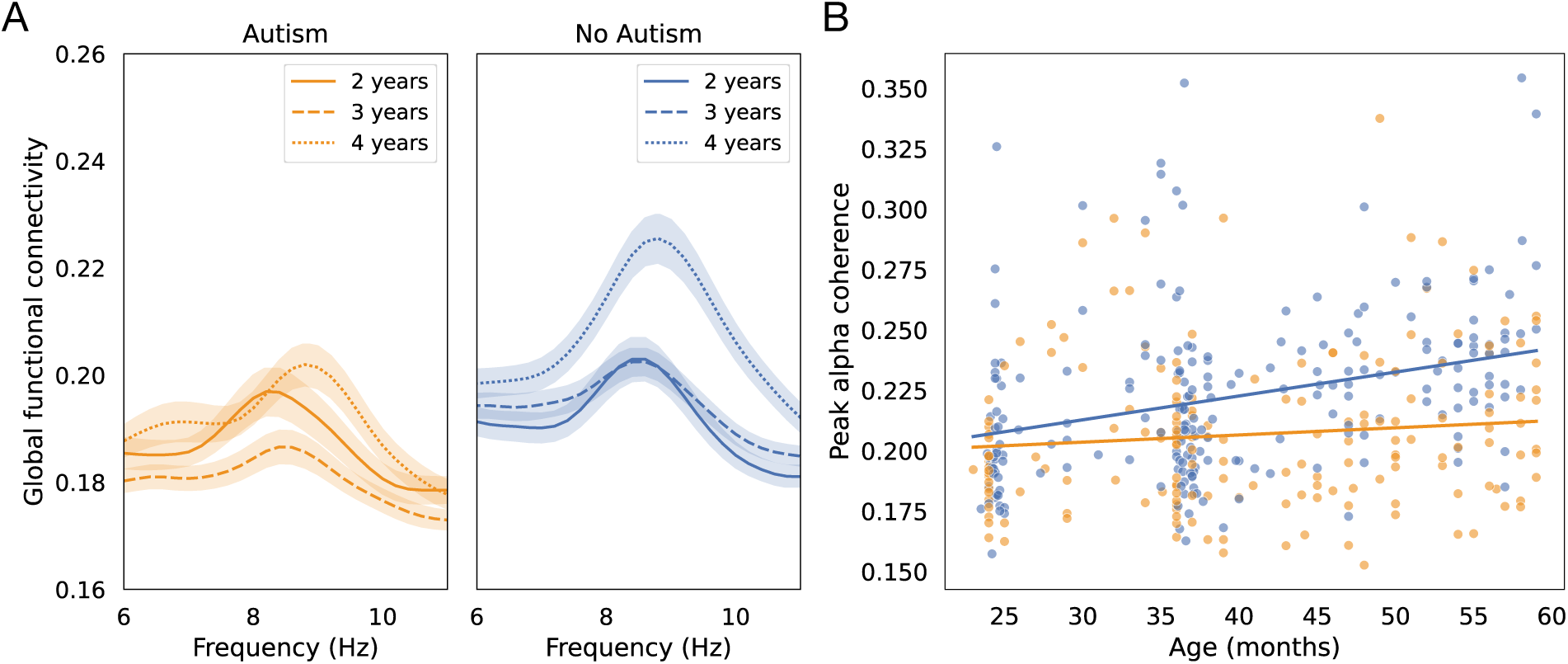
(A) Global functional connectivity stratified by group and age bin in the alpha frequency band. (B) Developmental trajectory of peak alpha coherence between 24 and 59 months in children with (orange) and without autism (blue).

### 3.3. EEG alpha coherence in autistic toddlers with and without language delay

The second aim of this study was to capture differences between children *within* the autism group. To do so, we assessed whether alterations in PAC observed in Aim 1 were driven by language delay status at 2, 3, and 4 years of age. Out of the 171 children diagnosed with autism, six did not complete the expressive language subscale of the MSEL. Therefore, language delay status could not be determined for these participants, and the following analysis comprised 165 measurements from autistic children aged 2 to 4 years of age (see Table S2 in the Supplement).

A significant fixed effect of age (β = 0.001, SE = 0.000, z = 2.35, p = .019) and a significant group x age interaction (β = −0.001, SE = 0.000, z = −2.09, p = .037) were found. As can be seen in Fig. 3, PAC significantly increased in autism-noLD (β = 0.001, SE = 0.000, z = 2.35, p = .019), while in autism-LD children, there was no evidence for a change in PAC over time (β = −0.000, SE = 0.000, z = −0.68, p = .495).

**Fig. 3.**
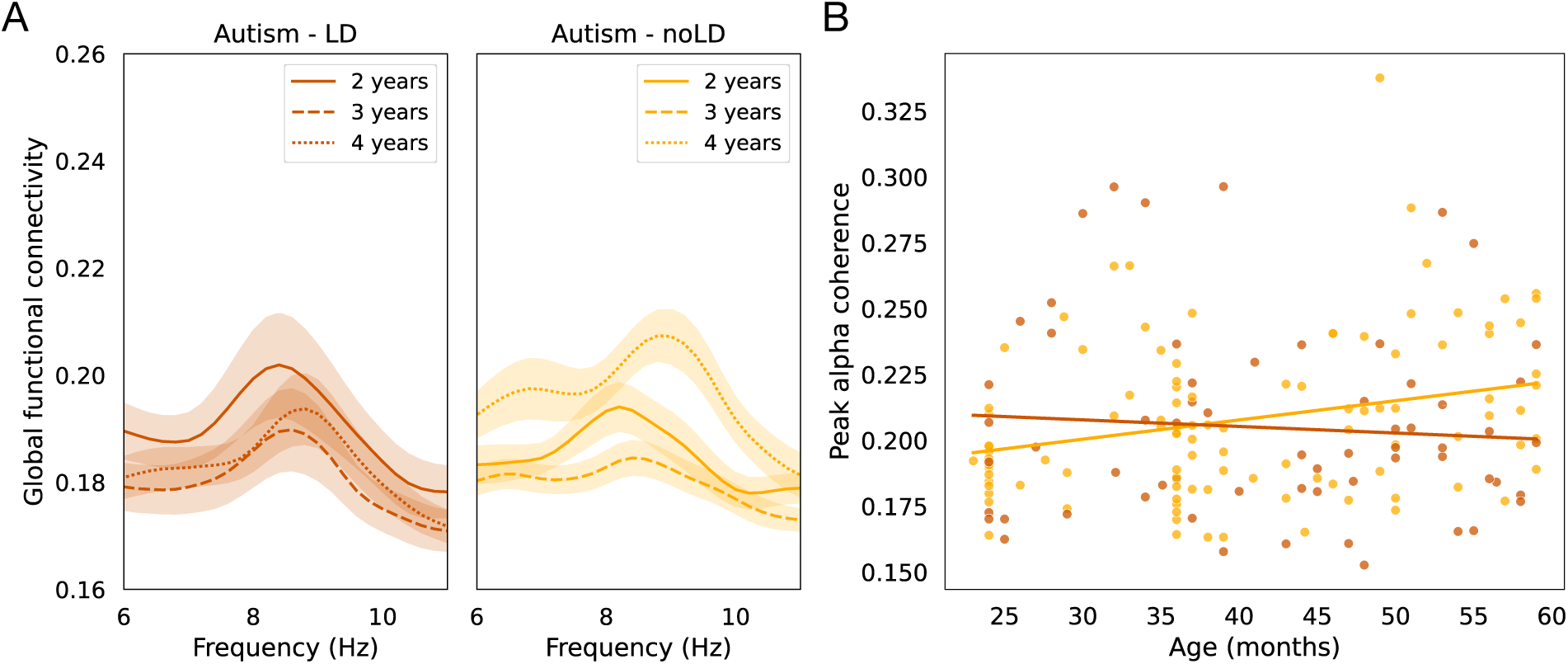
(A) Global functional connectivity stratified by group and age bin in the alpha frequency band. (B) Developmental trajectory of peak alpha coherence between 24 and 59 months in autistic children with LD (dark orange) and with noLD (light orange).

### 3.4. Exploratory analysis of peak alpha coherence and its association with language development

Given our above observation that the developmental trajectory of PAC differed between autistic children with and without language delay, we then explored the relationship between PAC and VDQ, and whether this link varied with age within the autism group. The analysis revealed evidence for an interaction between PAC and age (β = 13.18, 95% credible interval [1.96, 24.52]), suggesting that the association between PAC and VDQ becomes more positive with increasing age (Fig. 4.A; see Fig. S3 in the Supplement for non-modeled raw data associations). Although the interaction was positive, the estimated conditional effects were modest across much of the sampled age range and appeared strongest near the youngest and oldest participants, indicating substantial uncertainty of the observed effects across most of our observed age range (Fig. 4.B). In contrast, there was little evidence for a main effect of PAC (β = 2.41, 95% credible interval [−128.43, 128.08]) or age (β = 0.15, 95% credible interval [−0.22, 0.51]) in predicting VDQ.

**Fig. 4.**
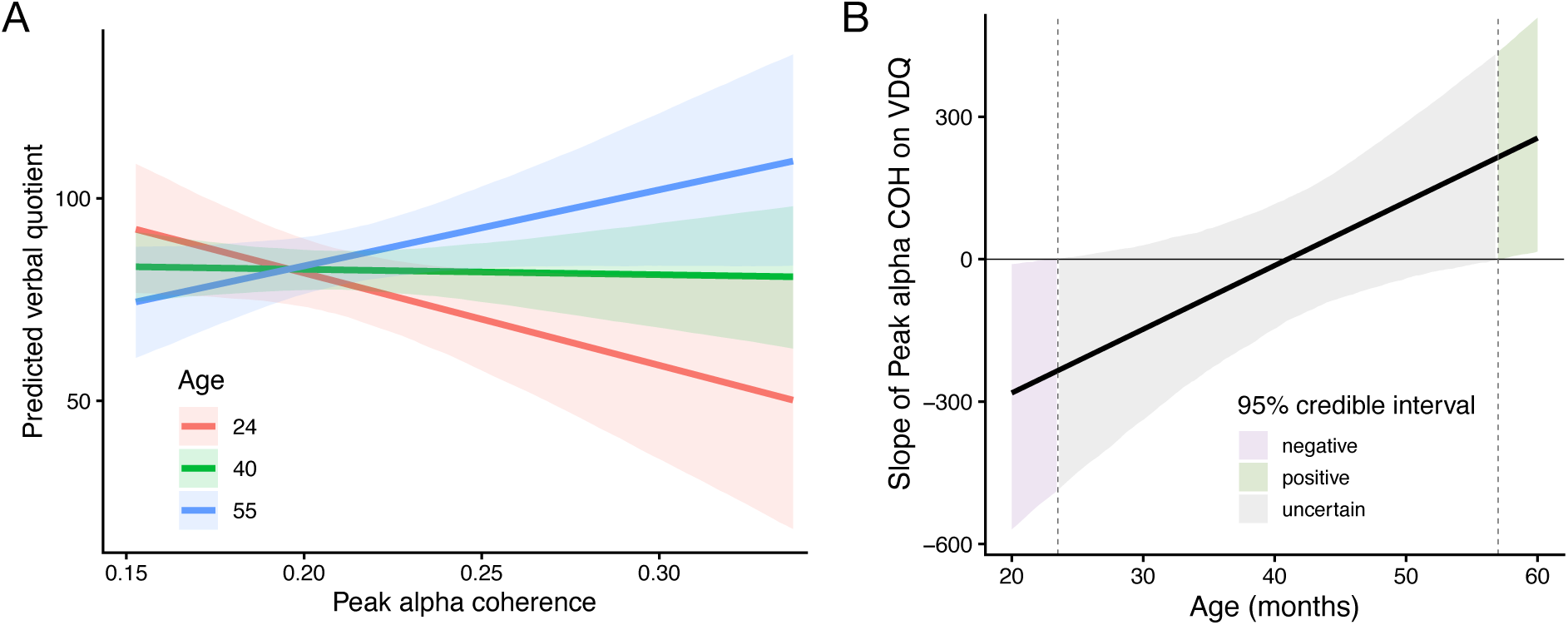
Age-dependent association between peak alpha coherence and verbal developmental quotient in autistic children. (A) Posterior estimated association between peak alpha coherence and predicted verbal developmental quotient at representative ages (24, 40, and 55 months) derived from the Bayesian mixed-effects model. (B) Posterior conditional slope estimates for the association between peak alpha coherence and verbal developmental quotient across the observed developmental range (20-60 months). The black line represents the posterior mean slope of peak alpha coherence across age, and shaded regions indicate 95% credible intervals. Colored regions denote ages at which the 95% credible interval for the conditional slope was below zero (negative), above zero (positive), or overlapped zero (uncertain).

## Discussion

The present study examined the developmental trajectory of PAC in (a) children with and without autism and (b) autistic children with and without language delay. The results supported our hypotheses: in neurotypical children, PAC increased between 2 and 4 years of age, whereas it did not significantly change with age in the overall autism group. Notably, this group difference appeared to be driven by autistic children with language delay, who showed an age-related decrease in PAC, in contrast to autistic children without language delay, who showed the expected age-related increase.

### 4.1. Neurodevelopmental trajectories in children with and without autism

The age-related increase in PAC observed in our neurotypical cohort is consistent with prior work and is generally interpreted as a progressive strengthening and stabilization of corticocortical connections through synaptic pruning, increased myelination, and experience-dependent plasticity (Anderson & Perone, 2018; Barry et al., 2004; Gmehlin et al., 2011). Alpha-band synchronization in particular is thought to support the coordination of activity across distributed brain networks involved in sensory integration, attention, and cognitive control (Palva & Palva, 2011; Sadaghiani & Kleinschmidt, 2016). Given this, the age-related increase in PAC observed in our neurotypical cohort between 2 and 4 years of age likely reflects the early strengthening and differentiation of large-scale functional networks.

In contrast, the absence of a comparable age-related PAC increase in autistic children may indicate atypical functional network maturation. This interpretation is supported by a recent study from our group that reported reduced global alpha coherence in autistic toddlers to be driven in part by diminished frontal connectivity, accompanied by reduced indices of network segregation (i.e., the pruning of connections between functional systems; Chung et al. (2026)). Moreover, prior functional MRI studies have shown that both network segregation and network integration (i.e., the strengthening of connections within functional systems) are reduced in older children and adolescents with autism compared to neurotypical controls, particularly in default-mode and higher-order visual regions (Rudie, Brown, et al., 2012; Rudie, Shehzad, et al., 2012). The attenuated growth of PAC observed in our preschool-aged group may thus signal that network-level disruptions emerge early in development in autism, which in turn may contribute to some of its core clinical features, including difficulties integrating complex social, sensory, and contextual information that rely on distributed networks.

### 4.2. Neurodevelopmental trajectories in autistic children with and without language delay

Beyond group-level differences in the developmental trajectories of PAC, our data also revealed distinct patterns within the autism cohort as a function of language ability. Whereas autistic children with no language delay exhibited an age-related increase in PAC from 2 to 4 years, those with language delay did not. This divergence in alpha coherence not only highlights substantial heterogeneity in the maturation of neural networks within autism but also suggests that individual differences in network strength may contribute to the variability in language abilities commonly seen in autism (Anderson et al., 2007; Delehanty et al., 2018).

To the best of our knowledge, there is little evidence on the relationship between alpha coherence and language abilities in children with autism (Gaudet et al., 2020). The alpha rhythm plays an important functional role in networks that support auditory processing and higher-order cognitive control, both of which are critical for language development (Palva & Palva, 2011; Sadaghiani & Kleinschmidt, 2016). The age-related increase in alpha coherence found in autistic children without language delay may thus reflect a more typical strengthening of functional connections within language-relevant circuits. This would also be consistent with previous evidence showing that stronger alpha coherence is associated with better semantic processing and word learning ability in neurotypical individuals (Huang et al., 2022; Rempe et al., 2022).

The absence of an increase in alpha coherence in autistic children with language delay, on the other hand, points to an altered trajectory of language-related network connectivity, possibly driven by delayed myelination or atypical synaptic pruning in neural systems supporting language development (Geschwind, 2011). Importantly, this divergence demonstrates that alpha coherence captures meaningful variability *within* the autism population, which is consistent with the view that alpha coherence patterns are generally altered in autism, but that its direction and spatial distribution vary with cognitive level and clinical phenotype (Schwartz et al., 2017). In turn, alpha coherence may serve as a sensitive biomarker of language development in autism, with potential utility for identifying children at risk for language delay. However, the opposite may also be true: differences in emerging language abilities may themselves shape the maturation of large-scale functional connectivity. Longitudinal studies with comprehensive language assessments are therefore needed to examine how alpha coherence and language abilities co-develop over time, and to clarify the directionality of this association.

### 4.3. The predictive value of peak alpha coherence for language ability

In a follow-up analysis, we examined whether PAC was associated with language ability (VDQ) and whether this relationship varied with age in children with autism. Although these findings should be interpreted with caution given their exploratory nature, the results revealed an interesting developmental pattern across the 2-to-4-year age range: higher PAC appeared to be associated with lower language ability at younger ages, whereas the opposite pattern emerged at older ages. This reversal in direction aligns with prior EEG and MEG studies showing that the relationship between neural measures and language abilities is not static across development and may differ depending on age (see Gaudet et al. (2020) for a review). Specifically, in neurotypical toddlers, coherence in the theta band has consistently predicted concurrent and later language outcomes (Kikuchi et al., 2011; Kühn-Popp et al., 2016; Mundy et al., 2003), while connectivity-language associations in other frequency bands, including alpha, beta, and gamma, appear to only become relevant in later childhood and adolescence (Doesburg et al., 2016; Doesburg et al., 2012). This age-dependent pattern of which frequency bands relate to language may reflect the broader maturation of neural oscillations, whereby slow-wave activity gradually gives way to faster frequencies across development (Uhlhaas et al., 2010). Our findings extend this picture to children with autism and suggest that even within the narrow 2-to-4-year window, the nature of the PAC-language relationship may itself be shifting. Future studies with larger samples and broader age ranges will be needed to carefully characterize the specific age-dependent shifts in functional connectivity patterns, thereby supporting a more nuanced understanding of brain-behavior associations across development.

### 4.4. Strengths and limitations

A key strength of the current study is its longitudinal design, which allowed us to examine developmental changes in PAC across early childhood and capture dynamic patterns of network organization in the preschool years. Moreover, by including both neurotypical children and a linguistically heterogeneous autism sample, we were able to not only assess group differences but also investigate how connectivity trajectories differ as a function of language ability within autism. Finally, the use of peak alpha coherence, rather than a fixed frequency band, offered a more individually tailored measure of alpha coherence that may better capture true maturational differences across children.

However, several limitations should also be acknowledged. The composition of the language-delayed subgroup in autism was not consistent across age bins, as some children caught up while others fell behind from one study visit to the next. Additionally, analyses were conducted at the sensor level rather than using source-localized signals, which limits the spatial specificity of the findings and the ability to attribute connectivity patterns to specific cortical regions. We therefore focused on whole-brain, or global, coherence patterns. Despite these limitations, the present study advances our understanding of the relationship between PAC and language development in young children with and without autism and highlights the importance of considering linguistic heterogeneity when examining neural markers within the autism spectrum.

## Data Availability

Consents obtained from human participants at Boston Children's Hospital prohibit sharing of identifiable and de-identified individual data without a data use agreement in place. Please contact the corresponding author with data requests. The code used for data analysis in this study will be made available on the Open Science Framework (OSF) upon publication.

## Acknowledgments

We would like to thank all children and families who participated in this research. We also thank Dr. Charles Nelson for sharing data from the Infant Sibling Project, and Dr. April Levin for sharing data from the Sensory Processing Adaptation Study. We are grateful to all the research staff across studies involved in participant recruitment, data collection, and database administration.

## Data and code availability

Consents obtained from human participants at our institution prohibit sharing of identifiable and de-identified individual data without a data use agreement in place. Please contact the corresponding author with data requests. The code used for data analysis in this study will be made available on the Open Science Framework (OSF) upon publication.

## Ethics approval

All four studies were approved by the Boston Children’s Hospital institutional review board and conducted in accordance with the Helsinki Declaration of 1975.

## Patient consent

A parent or legal guardian gave informed written consent before each child’s inclusion.

