## Supplemental Information for "The developmental trajectory of EEG alpha coherence in autistic toddlers with and without language delay"

**Supplement**

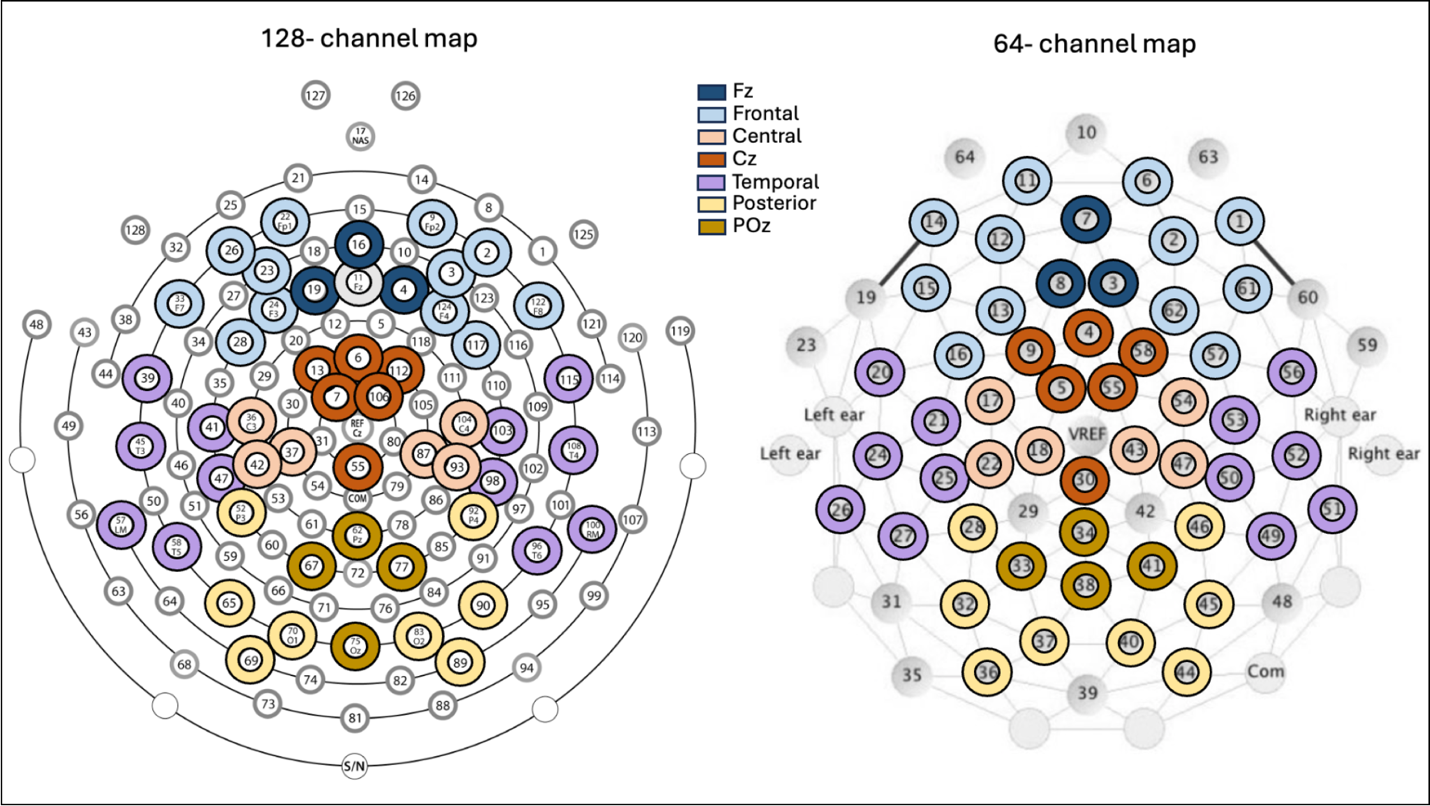

**Fig. S1**. Electrode selection. Topographic maps illustrating the original montages for the two EEG systems used across studies: a 128-channel system (left) and a 64-channel system (right). Colored circles indicate the 51 electrodes retained for analyses, which were selected to maximize spatial coverage while ensuring consistent electrode locations across both systems.

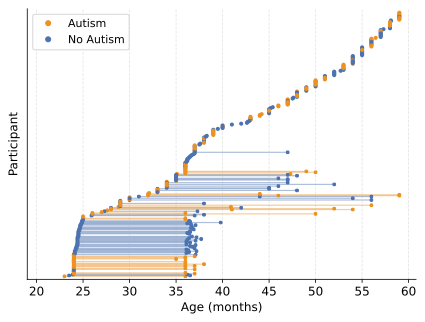
**Fig. S2.** Sample characteristics. Each row represents one participant, with dots indicating the age at time of assessment. Horizontal lines connected repeated measurements for participants with longitudinal data.

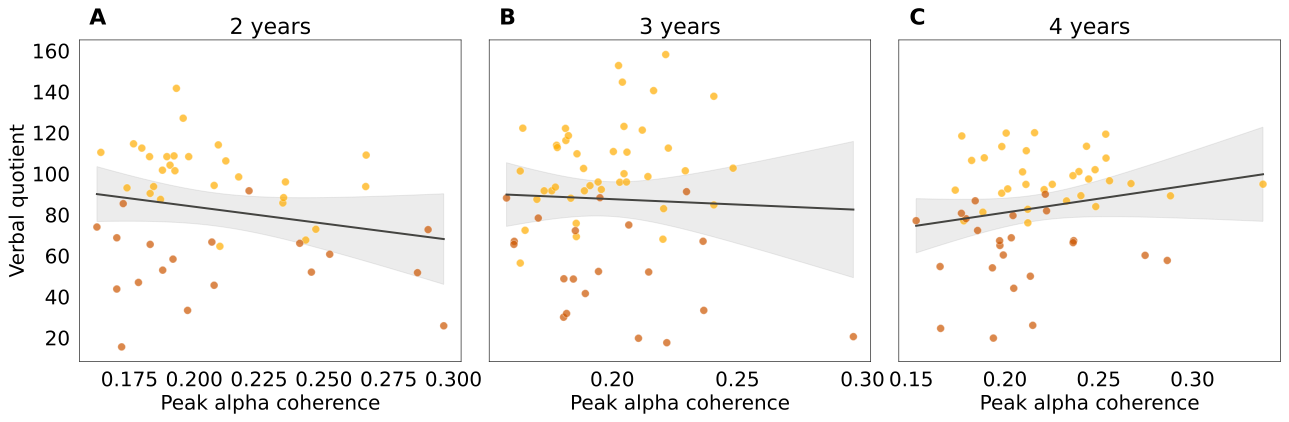

**Fig. S3.** Association between peak alpha coherence and verbal developmental quotient in autistic children at 2, 3, and 4 years of age. Raw data points are color-coded by language delay status.

**Table S1.** Number of participants from each study by age bin.

| **Study** | **N**  **2 years** | **N**  **3 years** | **N**  **4 years** |
| --- | --- | --- | --- |
| **BRIDGE** | | | |
| Autism | 5 | 3 | 0 |
| No autism | 0 | 0 | 0 |
| **IDEA** | | | |
| Autism | 21 | 16 | 43 |
| No autism | 22 | 15 | 37 |
| **ISP** | | | |
| Autism | 27 | 24 | 0 |
| No autism | 50 | 51 | 0 |
| **SPA** | | | |
| Autism | 0 | 23 | 13 |
| No autism | 0 | 29 | 20 |

**Table S2.** Sample size for each aim and analysis.

| **Analysis** | **N**  **total** | **N**  **no autism** | **N**  **autism-noLD** | **N**  **autism-LD** |
| --- | --- | --- | --- | --- |
| **Aim 1: autism vs. no autism** | 392 | 221 | 103 (6 unknown) | 62 |
| **Aim 2: autism-LD vs. autism-noLD** | 165 | - | 103 | 62 |
| **Exploratory analysis** | 164 | - | 164 | |
